# Association Between Parental Nativity and Mental Health Among U.S. Adolescents: Findings from the National Health Interview Survey, 2022–2024

**DOI:** 10.64898/2026.11.05.26367304

**Authors:** Sankhepo Ndhlovu, Zemu Liu, Lihua Li, Donglan Stacy Zhang

## Abstract

**Introduction:** Studies have shown a high prevalence of mental health problems among immigrants, but findings for their children are mixed. We examined the association between parental nativity and mental health among U.S. adolescents and assessed whether these associations varied by sex and race/ethnicity.

**Methods:** We analyzed pooled cross-sectional data from the 2022–2024 National Health Interview Survey, including 12,540 adolescents aged 12–17 years. The exposure was parental nativity. The outcomes were adolescents’ mental health, including anxiety, depression, behavioral difficulties, and peer relationships. Inverse probability of treatment weighting (IPTW) and nearest neighbor matching were used to control confounders. Survey-weighted logistic regression was used to estimate the association between parent nativity and each outcome. Subgroup analyses were conducted by sex and race/ethnicity.

**Results:** Compared with adolescents with U.S.-born parents, adolescents with at least one foreign-born parent had lower odds of anxiety (Odds Ratio [OR]=0.62, 95% confidence interval [CI]: 0.54– 0.71), depression (OR=0.71, 95% CI: 0.60–0.84), and behavioral difficulties (OR=0.56, 95% CI: 0.43–0.72), but not relationship difficulties (OR=0.97, 95% CI: 0.74–1.26), in the IPTW analysis. Findings from nearest-neighbor matching were consistent. In exploratory stratified analyses, inverse associations were observed among Hispanic adolescents and adolescents classified as other race/ethnicity.

**Conclusion:** Parental nativity was associated with adolescent mental health, with adolescents of at least one foreign-born parent showing lower odds of anxiety, depression, and behavioral difficulties. Future studies are needed to clarify the underlying mechanisms and identify protective factors that help inform interventions to promote adolescent mental health.

## Introduction

Adolescence (12–17 years) is a critical developmental period during which important physical, emotional, and social changes occur. These developmental transitions increase vulnerability to mental health conditions, including behavioral, emotional, psychological, and social problems, which may persist into adulthood [1]. In the United States (US), the prevalence of diagnosed mental health conditions among adolescents increased from 15.0% in 2016 to 20.3% in 2023, with more than 5.3 million adolescents having a current diagnosis of anxiety, depression, behavioral, or social problems [2]. These conditions are associated with a range of social, economic, and family-level factors [3], among which parental nativity has received increasing attention. Much of the literature examining children and adolescents wellbeing has documented the phenomenon of the immigrant paradox, whereby children of immigrants often exhibit more favorable health and developmental outcomes than their counterparts of US-born parents despite experiencing greater socioeconomic disadvantage and structural barriers [4-7]. Evidence supporting this paradox has been reported across a range of adolescents’ health outcomes, including mental health, behavioral functioning, and substance use [5,6,8,9]. However, findings have been mixed. While some studies report protective associations of immigrant family background [4,5], others have found no differences [10] or have observed variation across age at migration, duration of residence in the US, race and ethnicity, or other contextual factors [11,12].

Despite growing interest in parental nativity and adolescent mental health, important gaps remain. First, although the immigrant paradox has been widely examined across childhood and adolescence, relatively few studies have focused specifically on adolescents, a developmental period characterized by increasing independence and unique emotional, behavioral, and social challenges. Second, existing evidence is generally limited to single measures of mental health outcomes, such as depression [4], or have relied on regional cohort studies [9], limiting the generalizability of study findings. Third, prior studies have primarily relied on conventional multivariable regression approaches [4,11]. Propensity score methods provide an alternative framework for improving covariate balance between comparison groups and evaluating the robustness of observed associations [13-15]. Finally, fewer studies have evaluated whether associations between parental nativity and adolescent mental health differ across sociodemographic subgroups defined by race/ethnicity and sex.

To address these gaps, we examined associations between parental nativity and multiple domains of adolescent mental and behavioral health using nationally representative data from the National Health Interview Survey (NHIS). Guided by the immigrant paradox framework, which suggests that children of immigrant families may experience more favorable health outcomes despite socioeconomic disadvantage, we hypothesized that adolescents with foreign-born parent(s) had lower odds of adverse mental health outcomes than adolescents with US-born parents, partially due to family- and community-level protective factors, such as family cohesion, parental monitoring, social support, and cultural values. We further examined whether these associations varied by sex and race/ethnicity while accounting for demographic, socioeconomic, and household characteristics that may confound the relationship.

## Methods

### Data and Study Sample

We pooled cross-sectional data from the 2022–2024 NHIS Sample Child files. The NHIS is an annual, nationally representative survey of the civilian, noninstitutionalized US population conducted by the National Center for Health Statistics. Using a multistage, stratified, cluster sampling design, NHIS samples approximately 35,000 households each year and collects information on demographic characteristics, socioeconomic conditions, health status, health care access and utilization, and health-related behaviors. Since the 2019 redesign, NHIS has sampled approximately 35,000 households annually, with roughly 7,000–9,000 completed Sample Child interviews per year. The sample child component collects detailed information on 0-17 years old children’s health and well-being through interviews with a knowledgeable parent or guardian. In 2022, for example, NHIS included 7,464 completed Sample Child interviews, with a final Sample Child response rate of 45.8% [16-18].

Across the pooled 2022–2024 NHIS Sample Child files, 39,341 children participated in the Sample Child interview. For the present study, we restricted the analyses to adolescents aged 12– 17 years (N=14,644) to focus on a developmental period during which symptoms of depression, anxiety, behavioral difficulties, and social relationships become increasingly prevalent [1]. In total, 2,104 participants were excluded due to missing information on parental nativity, study outcomes, or covariates included in the propensity score model. The final analytic sample consisted of 12,540 adolescents, including 9,906 adolescents with US-born parents and 2,634 adolescents with at least one foreign-born parent (Figure S1).

### Exposure

The primary exposure was parental nativity, defined using the nativity of the responding parent or guardian interviewed in NHIS. This variable was coded binary and used to classify the study sample into two groups: adolescents whose interviewed parent or guardian was U.S.-born and those whose interviewed parent or guardian was foreign-born.

### Outcomes

We examined four domains of adolescent mental health, including frequent depression and anxiety symptoms, behavioral difficulties, and relationship difficulties.

Frequent depression and anxiety were derived from respondents’ answer to the survey questions: “How often does [sample child] seem very sad or depressed?” and “How often does [sample child] seem very anxious, nervous, or worried?”^16-18^ Consistent with prior NHIS-based research,[19] adolescents whose parent or guardian reported these symptoms occurring “daily” or “weekly” were classified as having frequent depression or anxiety symptoms, respectively. Responses of “monthly,” “a few times a year,” or “never” were classified as absence of frequent symptoms.

Behavioral difficulties were derived from respondents’ answer to the following survey questions: “Compared with children of the same age, does [sample child] have difficulty controlling his/her behavior?”, “Does [sample child] have difficulty concentrating on an activity that he/she enjoys doing?”, “Does [sample child] have difficulty accepting changes in his/her routine”. Adolescents were classified as having behavioral difficulties if the parent or guardian reported at least some difficulty on any of the three behavioral items. Those reported as having no difficulty on all three items were classified as not having behavioral difficulties. Relationship difficulty was based on the question “Does [sample child] have difficulty making friends?” Adolescents were classified as having behavioral or social difficulties if the parent or guardian reported the child had “some difficulty,” “a lot of difficulty,” or “cannot do at all” in the relevant domain [16-18]. Responses of “no difficulty” were classified as absence of any one of the three difficulties, consistent with prior literature [20].

### Covariates

Baseline covariates were selected a priori based on prior literature [21] Covariates included child age, sex (male or female); race/ethnicity (non-Hispanic White, non-Hispanic Black, Hispanic, and Other, which included non-Hispanic Asian, non-Hispanic American Indian/Alaska Native, and non-Hispanic multiracial adolescents); parent or guardian educational attainment (less than high school, high school graduate or equivalent, some college or associate degree, and bachelor’s degree or higher), parent or guardian employment status (employed vs. not employed); household income-to-poverty ratio (<100%, 100%–199%, 200%–399%, and ≥400% of the federal poverty level); and household structure (two-parent household vs. single-parent/other household).

### Statistical Analysis

To reduce confounding due to demographic, socioeconomic, and household differences between adolescents with foreign-born and US-born parents, we used inverse probability of treatment weighting (IPTW) for primary analyses. Stabilized weights were derived from propensity scores estimated via survey-weighted logistic regression [14]. The model used to estimate the propensity scores included all the baseline covariates. Baseline characteristics were compared before and after weighting, and covariate balance was assessed using standardized mean differences (SMD) with values less than 0.10 indicating adequate balance [15]. To estimate the population-level association between parental nativity and each outcome, we fitted generalized linear models with a quasibinomial family with a logit link function, weighted by the product of the IPTW and NHIS sampling weights [13]. Quasibinomial models were used to account for potential overdispersion in survey-weighted binary outcomes. Because the analyses pooled data from three survey years (2022–2024), the sampling weights were divided by the number of years pooled, as recommended by NHIS method guide [22]. We assessed whether associations varied across demographic subgroups by conducting stratified IPTW analyses by sex and race/ethnicity.

As a sensitivity analysis, we performed nearest-neighbor matching to create comparable exposure groups. Nearest-neighbor matching was based on propensity score distance and was performed without replacement using 1 up to 3 matching ratio, with exact matching on sex, race/ethnicity, parental education, employment status, poverty status, and family structure.

Adolescents were additionally matched on age with caliper width of +/- 1 year. This resulted in a final matched sample of 2,206 adolescents with a foreign-born parent and 4,730 adolescents with U.S.-born parents. We then applied the same generalized linear models with a quasibinomial link function to the matched sample, while weighted by the NHIS sampling weights.

The analyses were conducted using R version 4.5.2. All analyses incorporated the complex survey design, including sampling weights, primary sampling units and stratification variables. Results are presented as odds ratios (ORs) with 95% confidence intervals (CIs). Statistical significance was defined as a two-sided p-value < 0.05.

## Results

### Characteristics of study population

Before weighting, compared to adolescents with US-born parents, those with foreign-born parents were more likely to be Hispanic (43.5% vs. 21.3%), have parents with less than a high school education (22.9% vs. 6.3%), and live below the federal poverty level (16.1% vs. 10.9%). After IPTW, covariate balance improved substantially, with all SMD below 0.10 (Table 1). Adequate covariate balance was also achieved after matching, with all standardized mean differences below 0.10 (Table S1).

**Table 1:**
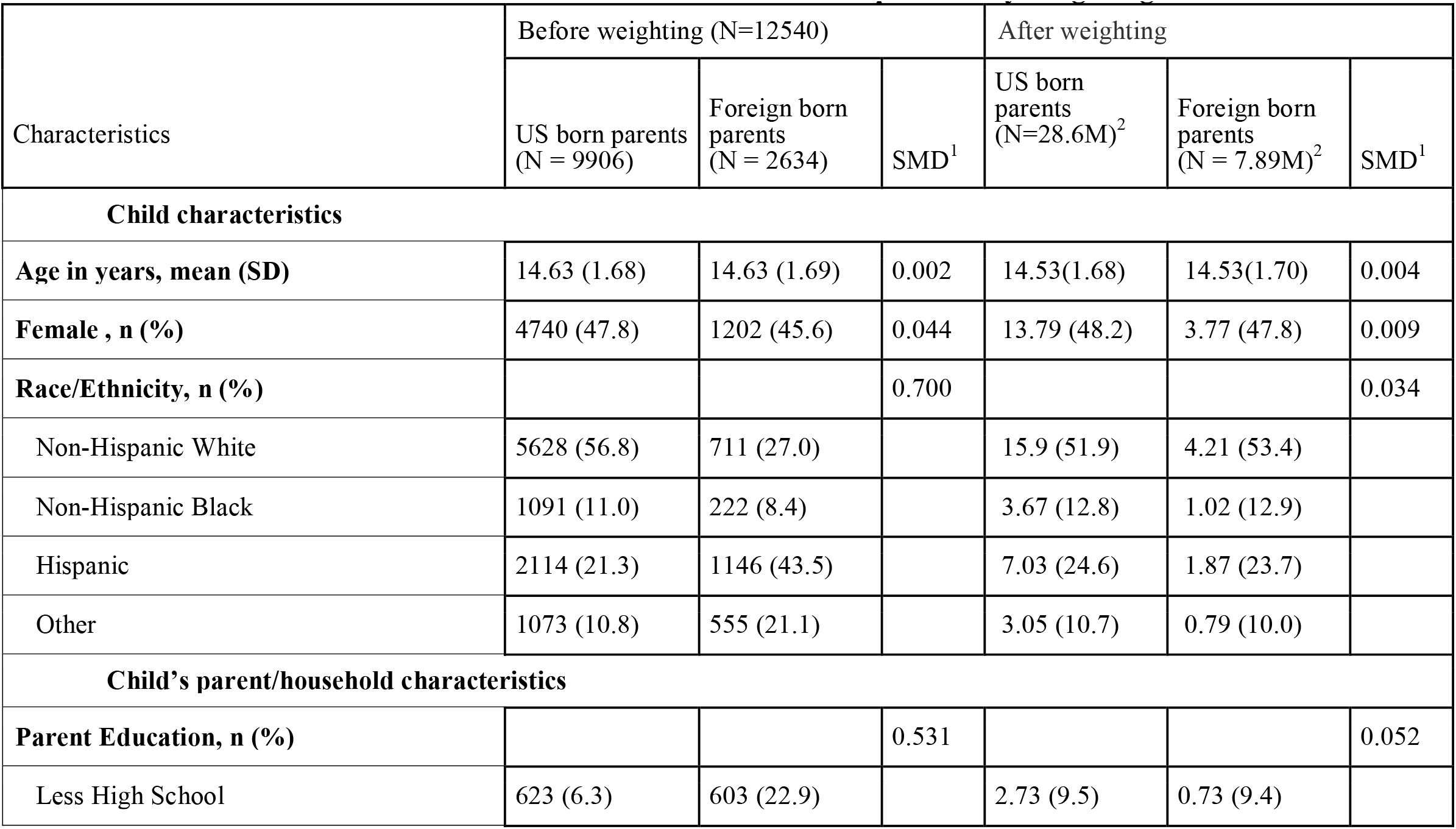
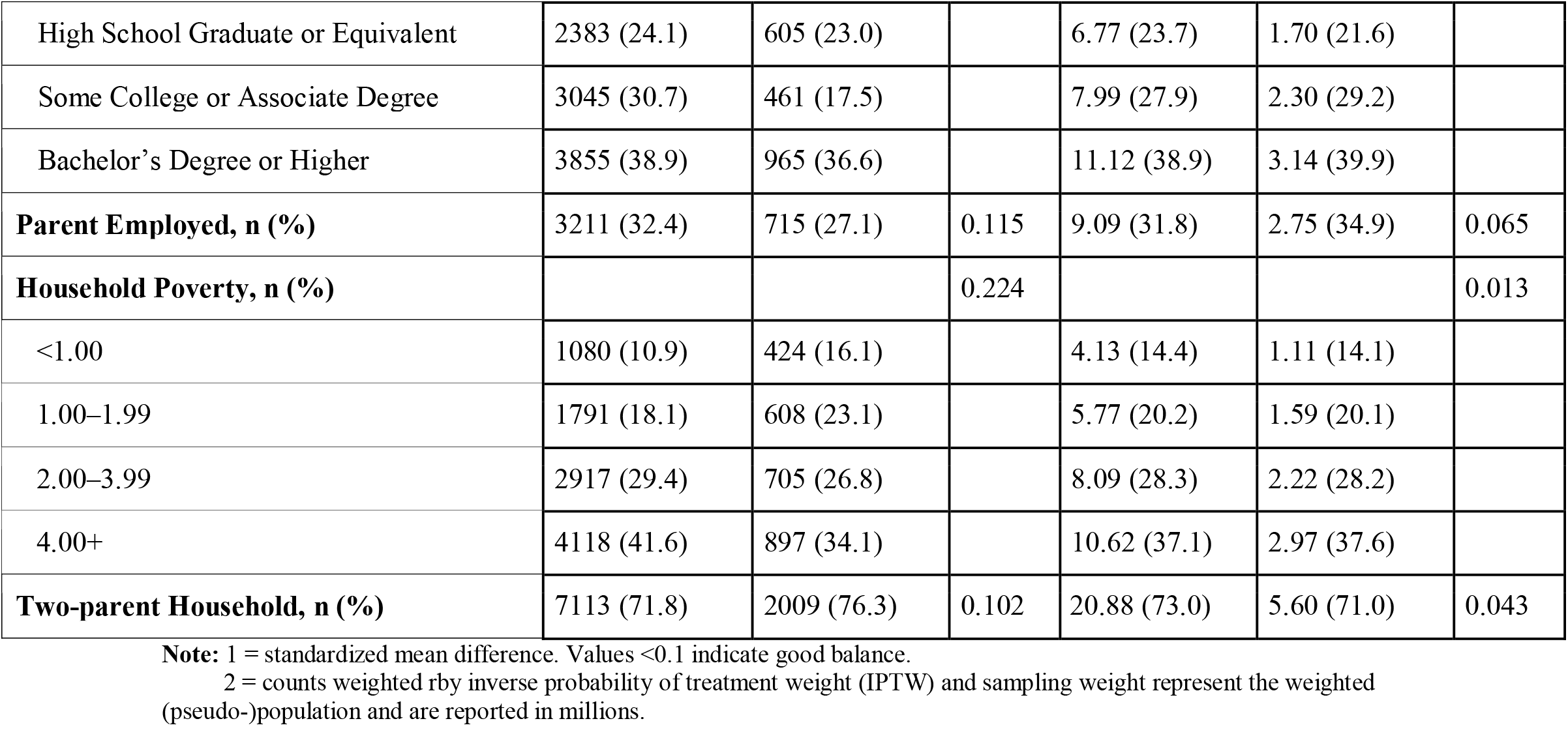
Baseline characteristics of adolescents aged 12–17 by parental nativity, with standardized mean differences before and after inverse probability weighting.

### Association Between Parental Nativity and Adolescent Mental and Behavioral Health Outcomes

Adverse mental health outcomes were generally less common among adolescents with at least one foreign-born parent than among adolescents with US-born parents. Among adolescents with US-born parents, the weighted prevalence of frequent anxiety, depression, and behavioral difficulties was 33.2%, 16.7%, and 6.7%, respectively, compared with 23.6%, 12.4%, and 4.3%, respectively, among adolescents with at least one foreign-born parent. The weighted prevalence of relationship difficulties was similar between the two groups (3.3% vs. 3.4%) (Figure 1).

**Fig. 1.**
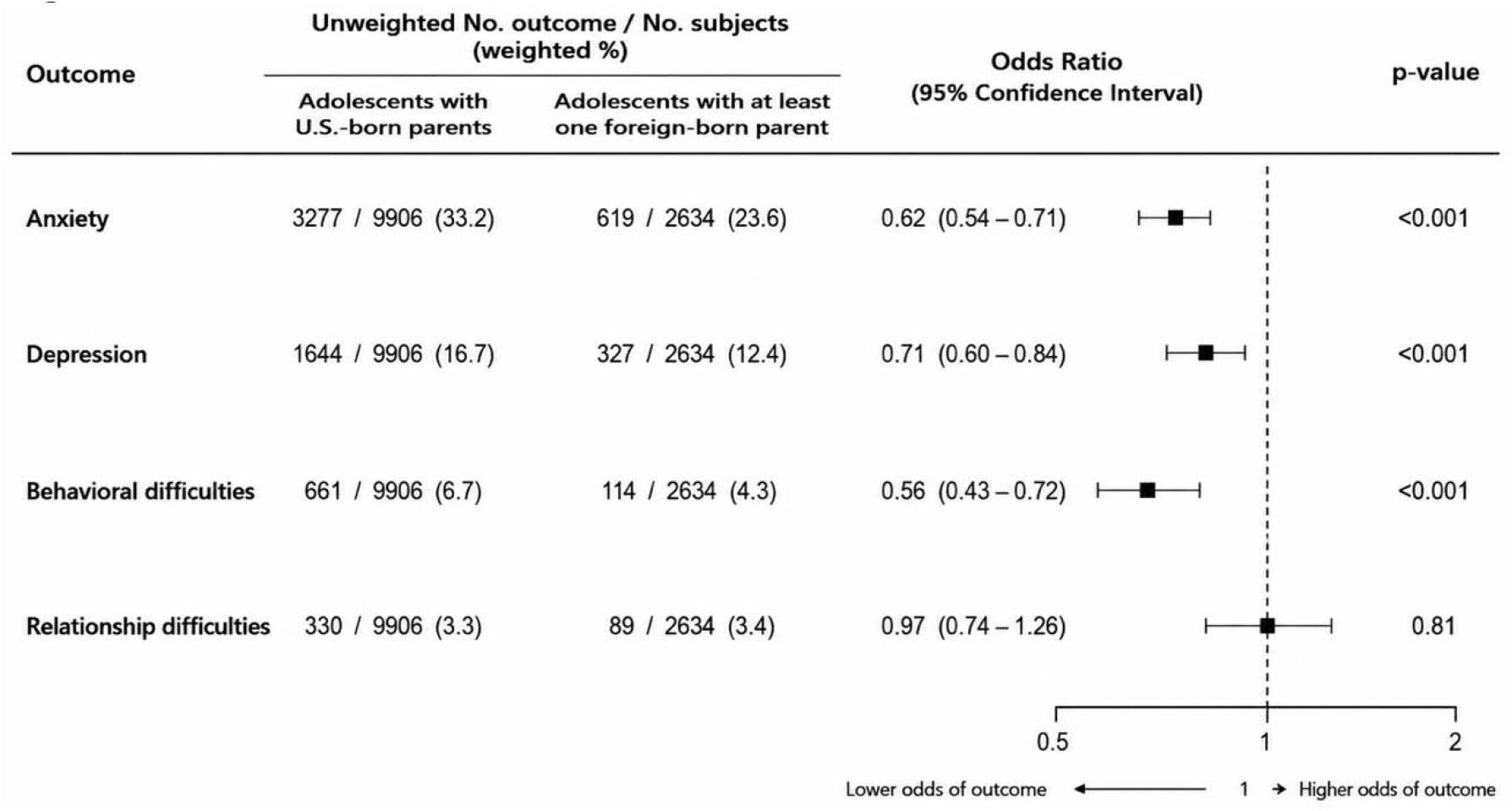

Estimated association between parental nativity and mental health outcomes among adolescents aged 12–17, using inverse probability of treatment weighting analyses.

Results from IPTW analyses show that adolescents with at least one foreign-born parent had lower odds of anxiety (OR = 0.62; 95% CI: 0.54–0.71), depression (OR = 0.71; 95% CI: 0.60– 0.84), and behavioral difficulties (OR = 0.56; 95% CI: 0.43–0.72), compared to adolescents with US-born parents. No statistically significant association was observed for relationship difficulties (OR = 0.97; 95% CI: 0.74–1.26) (Figure 1). Results from the nearest-neighbor matching analysis were consistent with the IPTW findings, with estimated ORs of 0.68 (95% CI:0.61–0.77), 0.71 (95% CI:0.62–0.83) and 0.70 (95% CI:0.54–0.89) for frequent anxiety symptoms, frequent depression symptoms and behavioral difficulties respectively. Similarly, no significant association was observed for relationship difficulties (OR=1.05; 95% CI:0.79–1.40). (Figure S2).

### Association Between Parental Nativity and Adolescent Mental Health and Behavioral Difficulties Stratified by Sex and Race/Ethnicity

Among male adolescents with US-born parents, the weighted prevalence of anxiety, depression, behavioral difficulties, and relationship difficulties was 27.6%, 11.7%, 7.2%, and 3.3%, respectively, while this prevalence was 19.6%, 8.7%, 5.0%, and 3.3%, respectively among those with at least one foreign-born parent. Having at least one foreign-born parent was associated with lower odds of anxiety (OR = 0.63; 95% CI, 0.53–0.75), depression (OR = 0.71; 95% CI, 0.56–0.91), and behavioral difficulties (OR = 0.58; 95% CI, 0.42–0.79), whereas no significant association was observed for relationship difficulties (OR = 0.87; 95% CI, 0.60–1.25). Similar patterns were observed among female adolescents. The weighted prevalence of anxiety, depression, behavioral difficulties, and relationship difficulties was 39.0%, 21.9%, 6.1%, and 3.4%, respectively, among adolescents with US-born parents, compared with 28.2%, 16.8%, 3.5%, and 3.5%, respectively, among those with at least one foreign-born parent. Similar associations were also observed among female adolescents for anxiety (OR = 0.60; 95% CI, 0.50–0.74), depression (OR = 0.72; 95% CI, 0.58–0.90), and behavioral difficulties (OR = 0.52; 95% CI, 0.36–0.77), while no significant association was observed for relationship difficulties (OR = 1.09; 95% CI, 0.74–1.63) (Figure 2).

**Fig. 2.**
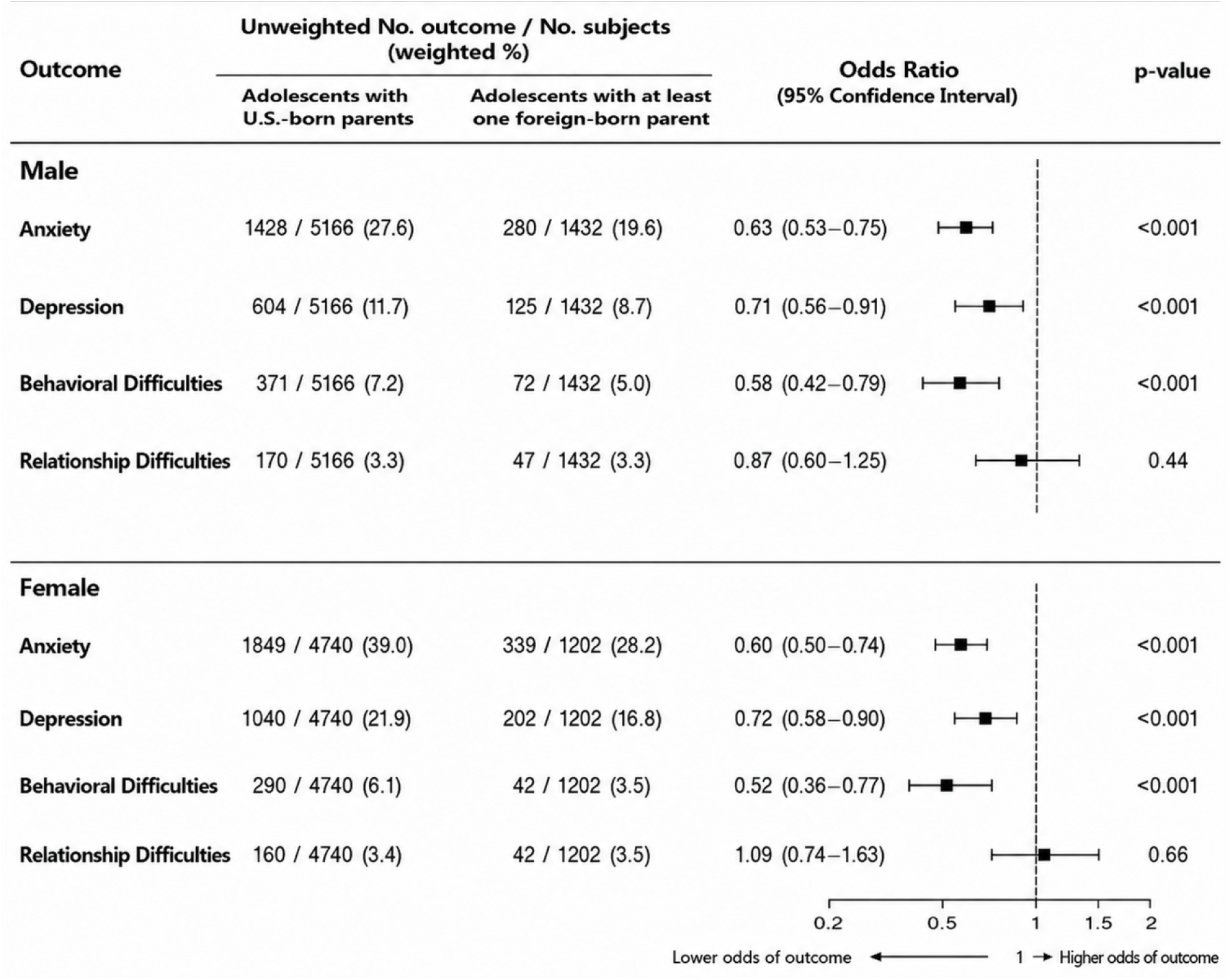
Association Between Parental Nativity and Adolescent Mental Health Outcomes Stratified by Sex

Across racial and ethnic subgroups, the weighted prevalence of frequent anxiety, frequent depression, behavioral difficulties, and relationship difficulties was again lower among adolescents with at least one foreign-born parent than among those with US-born parents. Among non-Hispanic White adolescents, the corresponding prevalences were 39.5%, 18.7%, 7.9%, and 4.0% versus 38.1%, 17.0%, 6.2%, and 4.6%. Among non-Hispanic Black adolescents, the prevalences were 21.2%, 11.7%, 4.8%, and 1.5% versus 16.7%, 11.7%, 3.6%, and 3.2%.

Among Hispanic adolescents, the prevalences were 25.2%, 14.0%, 5.3%, 3.0% versus 17.5%, 10.8%, 3.8% and 2.9%. And these corresponding prevalences were 27.2%, 15.7%, 5.0%, and 2.3% versus 19.8%, 10.1%, 3.4%, and 2.9%, among other racial or ethnic adolescents. Having at least one foreign-born parent was associated with lower odds of frequent anxiety and depression among Hispanic adolescents (anxiety: OR=0.68; 95% CI:0.54–0.86; depression: OR, 0.71; 95% CI, 0.53–0.96) and other racial and ethnic adolescents (anxiety: OR=0.60; 95% CI:0.43–0.80; depression: OR=0.65; 95% CI:0.45–0.95). Lower odds of behavioral difficulties were only observed among Hispanic adolescents (OR=0.59; 95% CI:0.38–0.92) (Figure 3).

**Fig. 3.**
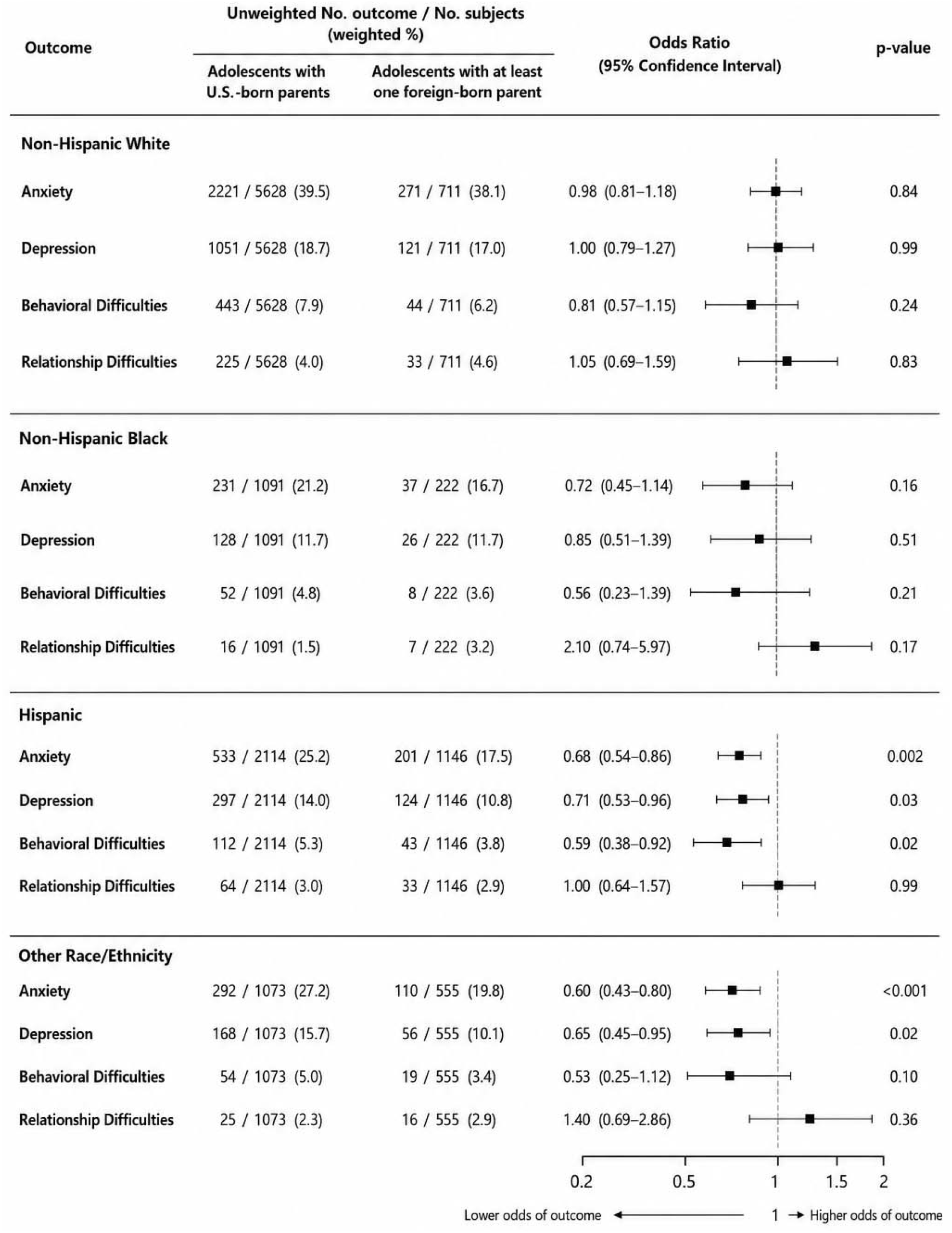
Association Between Parental Nativity and Adolescent Mental Health Outcomes Stratified by Race/Ethnicity

## Discussion

In this study, we conducted both IPTW and nearest neighbor matching analyses of a nationally representative sample of US adolescents aged 12-17 years. We found that those with at least one foreign-born parent had lower odds of anxiety, depression, and behavioral difficulties than their peers with US-born parents. This is consistent with the immigrant paradox described in prior studies where lower prevalence of psychiatric disorders, emotional problems, and behavioral difficulties were reported among children from immigrant families compared with children of US-born parents [23-25].

Several mechanisms may explain these findings. Prior studies suggest that immigrant families may benefit from protective social and cultural resources, including stronger family cohesion, greater family obligations, closer parental monitoring, collectivist cultural norms, and supportive social networks [5,25-30]. Such factors may foster resilience by strengthening parent-child relationships, promoting emotional support, and encouraging adaptive coping strategies during adolescence. As a result, these influences may buffer adolescents from depression, anxiety, and behavioral difficulties despite exposure to socioeconomic disadvantages. Identifying and strengthening these protective factors may inform the development of culturally responsive mental health interventions and prevention strategies.

However, we did not observe an association between parental nativity and relationship difficulties between the two groups of adolescents. This finding suggests that protective effects may not extend uniformly across all components of mental health. It may be more evident for internalizing symptoms and behavioral problems than for peer relationships and social interactions. One possible explanation is that peer relationships are influenced by factors beyond the family context. Although immigrant families may provide strong internal protective resources, they may also face challenges such as language barriers, cultural differences, residential instability, or unfamiliarity with community resources, all of which may reduce opportunities for social integration and peer engagement [28,31].

We also found that the association between parental nativity and mental health was not uniform across racial and ethnic groups. In exploratory stratified analyses, associations appeared stronger among Hispanic adolescents and adolescents classified as other race/ethnicity, although formal interaction testing was not performed, whereas estimates among non-Hispanic White and non-Hispanic Black adolescents were generally closer to the null or less precise. This is not surprising, given immigrant families are highly heterogeneous, and these protective factors may vary substantially according to country of origin, migration experiences, acculturation patterns, and family circumstances [8,23,25,31,32]. This finding suggests that immigrant family background, along with cultural and social factors, also shape adolescent mental health outcomes. Because formal interaction tests were not conducted, these findings should not be interpreted as evidence of effect modification. Future studies should explore mechanisms, such as family cohesion, acculturation, social support, that may explain variation in mental health trajectories among children, particularly children from immigrant families across racial and ethnic groups.

Our study has several strengths. First, it measures adolescent mental health across multiple domains including anxiety, depression, behavioral difficulties, and relationship difficulties.

Second, it leverages a recent nationally representative sample of U.S. adolescents and accounts for complex survey design, enabling study findings to be generalizable to the national adolescent population. Third, the use of both IPTW and nearest-neighbor matching methods yielded consistent findings increasing confidence that the observed associations were not sensitive to the choice of analytic approaches.

Several limitations should be considered when interpreting these findings. First, the NHIS is cross-sectional, precluding assessment of temporal ordering and causal inferences. Second, all mental health outcomes were based on parent or guardian reports rather than adolescent self-report, potentially introducing reporting bias. Parents or guardians may differ in their ability to recognize, interpret, or report symptoms of anxiety, depression, and behavioral difficulties, and these differences may vary across cultural contexts because of stigma, help-seeking norms, or perceptions of mental health. Consequently, some of the observed associations may reflect differences in symptoms reporting rather than true differences in adolescent mental health. Third, parental nativity was determined using the interviewed parent or guardian only, because information on the nativity of the nonresponding parent was unavailable. As a result, our measure reflects the nativity of the responding parent/guardian rather than both parents, potentially leading to exposure misclassification and preventing use from distinguishing adolescents with one foreign-born parent from those with two foreign-born parents. In addition, the identity of the responding parent or guardian may have influenced outcome reporting. Future studies incorporating adolescent self-report and nativity information from both parents could provide a more comprehensive assessment. Finally, NHIS does not capture several immigration-related factors that may be relevant to adolescent mental health, including parental duration of residence in the United States, documentation status, acculturation, language use, social network and support, and experiences of discrimination.

## Conclusion

We found that adolescents with at least one foreign-born parent had lower odds of anxiety, depression, and behavioral difficulties than adolescents with US-born parents, suggesting that parental nativity may be an important social determinant of youth mental health. Future studies, particularly with adolescent self-report measures, are needed to validate our findings and identify the mechanisms underlying these associations.

## New Contributions to the Literature

Prior literature has established that children of foreign-born parents often experience better health outcomes than children of U.S.-born parents despite greater socioeconomic disadvantage. However, to our knowledge, we are among the first to provide evidence on mental health across multiple domains using a nationally representative sample of U.S. adolescents. We found that adolescents with at least one foreign-born parent had lower odds of anxiety, depression, and behavioral difficulties. Understanding protective factors within immigrant families may help inform targeted intervention and prevention strategies for promoting mental health well-being among U.S. adolescents.

## Data Availability

All data produced are available online at
CDC National Center for Health Statistics
National Health Interview Survey
https://www.cdc.gov/nchs/nhis/documentation/index.html

## References

1. Centers for Disease Control and Prevention. About Mental Health. Updated May 19, 2026. Accessed August 6, 2026, https://www.cdc.gov/mental-health/about/index.html

2. Health Resources and Services Administration’s Maternal and Child Health Bureau. National Survey of Children’s Health: Adolescent Mental and Behavioral Health, 2023 Accessed August 7, 2026, https://mchb.hrsa.gov/sites/default/files/mchb/data-research/nsch-data-brief-adolescent-mental-behavioral-health-2023.pdf

3. Zarei K, Kahle L, Buckman DW, et al. Parent-Child Nativity, Race, Ethnicity, and Common Mental Health Conditions Among United States Children and Adolescents. J Pediatr. Dec 2023;263:113618. doi:10.1016/j.jpeds.2023.113618

4. Harker K. Immigrant Generation, Assimilation, and Adolescent Psychological Well-Being. Social Forces. 2001;79(3):969–1004. doi:10.1353/sof.2001.0010

5. Marks AK, Ejesi K, Garcías Coll C. Understanding the U.S. Immigrant Paradox in Childhood and Adolescence. Child Development Perspectives. 2014;8(2):59–64. doi:10.1111/cdep.12071

6. Salas-Wright CP, Vaughn MG, Schwartz SJ, Cordova D. An “immigrant paradox” for adolescent externalizing behavior? Evidence from a national sample. Soc Psychiatry Psychiatr Epidemiol. Jan 2016;51(1):27–37. doi:10.1007/s00127-015-1115-1

7. Coll CG, Marks AK. The immigrant paradox in children and adolescents: Is becoming American a developmental risk? 2012.

8. Elshahat S, Moffat T, Newbold KB. Understanding the Healthy Immigrant Effect in the Context of Mental Health Challenges: A Systematic Critical Review. J Immigr Minor Health. Dec 2022;24(6):1564–1579. doi:10.1007/s10903-021-01313-5

9. Fuligni AJ. Authority, autonomy, and parent-adolescent conflict and cohesion: a study of adolescents from Mexican, Chinese, Filipino, and European backgrounds. Dev Psychol. Jul 1998;34(4):782–92. doi:10.1037//0012-1649.34.4.782

10. Feliciano C, Lanuza YR. An Immigrant Paradox? Contextual Attainment and Intergenerational Educational Mobility. American Sociological Review. 2017;82(1):211–241. doi:10.1177/0003122416684777

11. Filion N, Fenelon A, Boudreaux M. Immigration, citizenship, and the mental health of adolescents. PLoS One. 2018;13(5):e0196859. doi:10.1371/journal.pone.0196859

12. Teruya SA, Bazargan-Hejazi S. The Immigrant and Hispanic Paradoxes: A Systematic Review of Their Predictions and Effects. Hisp J Behav Sci. Sep 5 2013;35(4):486–509. doi:10.1177/0739986313499004

13. Applied Survey Data Analysis, Second Edition. 2017.

14. Rosenbaum PR, Rubin DB. The central role of the propensity score in observational studies for causal effects. Biometrika. 1983;70(1):41–55. doi:10.1093/biomet/70.1.41

15. Austin PC. Balance diagnostics for comparing the distribution of baseline covariates between treatment groups in propensity_Jscore matched samples. Statistics in Medicine. 2009;28(25):3083–3107. doi:10.1002/sim.3697

16. Division of Health Interview Statistics, National Center for Health Statistics, Centers for Disease Control and Prevention, U.S. Department of Health and Human Services. National Health Interview Survey: 2022 Survey Description. Centers for Disease Control and PreventionU.S. Department of Health and Human Services. Accessed August 7, 2026, https://ftp.cdc.gov/pub/Health_Statistics/NCHS/Dataset_Documentation/NHIS/2022/srvydesc-508.pdf

17. Division of Health Interview Statistics, National Center for Health Statistics, Centers for Disease Control and Prevention, U.S. Department of Health and Human Services. National Health Interview Survey: 2023 Survey Description. Centers for Disease Control and Prevention, U.S. Department of Health and Human Services. Accessed August 7, 2026, https://ftp.cdc.gov/pub/Health_Statistics/NCHS/Dataset_Documentation/NHIS/2023/srvydesc-508.pdf

18. Division of Health Interview Statistics, National Center for Health Statistics, Centers for Disease Control and Prevention, U.S. Department of Health and Human Services. National Health Interview Survey: 2024 Survey Description. Centers for Disease Control and Prevention, U.S. Department of Health and Human Services. Accessed August 7, 2026, https://ftp.cdc.gov/pub/Health_Statistics/NCHS/Dataset_Documentation/NHIS/2024/srvydesc-508.pdf

19. Zablotsky B, Ng AE, Black LI, Blumberg SJ. Diagnosed Developmental Disabilities in Children Aged 3-17 Years: United States, 2019-2021. NCHS Data Brief. Jul 2023;(473):1–8.

20. Weeks JD, Madans JH, Black LI, Elgaddal N, Zablotsky B. Difficulties in Functioning Among Children in the United States: 2021-2023. Natl Health Stat Report. Sep 9 2025;(218):1. doi:10.15620/cdc/174615

21. Hernán MA, Robins JM. Causal inference: what if. First edition. ed. Taylor and Francis,; 2020:1 online resource.

22. Rivera Drew JA. Guidance for Pooling Multiple Years of NHIS Data. Accessed August 7, 2026, https://blog.popdata.org/pooling-years-of-nhis-data/

23. Gadermann AM, Alonso J, Vilagut G, Zaslavsky AM, Kessler RC. Comorbidity and disease burden in the National Comorbidity Survey Replication (NCS-R). Depress Anxiety. Sep 2012;29(9):797–806. doi:10.1002/da.21924

24. Singh GK, Yu SM, Siahpush M, Kogan MD. High levels of physical inactivity and sedentary behaviors among US immigrant children and adolescents. Arch Pediatr Adolesc Med. Aug 2008;162(8):756–63. doi:10.1001/archpedi.162.8.756

25. Alegria M, Alvarez K, DiMarzio K. Immigration and Mental Health. Curr Epidemiol Rep. Jun 2017;4(2):145–155. doi:10.1007/s40471-017-0111-2

26. Marsiglia FF, Parsai M, Kulis S, Southwest Interdisciplinary Research C. Effects of Familism and Family Cohesion on Problem Behaviors among Adolescents in Mexican Immigrant Families in the Southwest U.S. J Ethn Cult Divers Soc Work. Jul 1 2009;18(3):203–220. doi:10.1080/15313200903070965

27. Bacong AM, Menjivar C. Recasting the Immigrant Health Paradox Through Intersections of Legal Status and Race. J Immigr Minor Health. Oct 2021;23(5):1092–1104. doi:10.1007/s10903-021-01162-2

28. Lerias D, Ziaian T, Arthur N, Augoustinos M, Pir T, Miller E. The Mental Health of Refugee and Migrant Youth after Settlement: Outcomes of a Multinational Study. Community Ment Health J. Oct 2025;61(7):1334–1367. doi:10.1007/s10597-025-01474-9

29. Bunn M, Betancourt T. Strengthening mental health support services for refugee children resettled in the US. Child Evidence Briefs. 2022;16

30. Shah S, Choi M, Miller M, Halgunseth LC, van Schaik SDM, Brenick A. Family cohesion and school belongingness: Protective factors for immigrant youth against bias-based bullying. New Dir Child Adolesc Dev. May 2021;2021(177):199–217. doi:10.1002/cad.20410

31. Roche KM, White RMB, Partovi R, Vaquera E, Little TD. US Immigration Policy Stressors and Latinx Youth Mental Health. JAMA Pediatr. Jul 1 2024;178(7):669–677. doi:10.1001/jamapediatrics.2024.1153

32. Dimitrova R, Chasiotis A, van de Vijver F. Adjustment outcomes of immigrant children and youth in Europe: A meta-analysis. European Psychologist. 2016;21(2):150–162. doi:10.1027/1016-9040/a000246

