## Supplementary Material for "Association Between Parental Nativity and Mental Health Among U.S. Adolescents: Findings from the National Health Interview Survey, 2022–2024"

**Table S1: Baseline characteristics of adolescents aged 12–17 by parental nativity, with standardized mean differences before and after matching**

| Characteristics | Before Matching (N=12540) | | | After Matching (N=6933) | | |
|---|---|---|---|---|---|---|
| | US born parents (N = 9,906) | Foreign born parents (N = 2,634) | SMD[1] | US born parents (N = 4,727) | Foreign born parents (N = ,2206) | SMD[1] |
| ***Child characteristics*** | | | | | | |
| **Age in years (mean, SD)** | 14.63 (1.68) | 14.63 (1.69) | 0.002 | 14.69 (1.65) | 14.56 (1.67) | 0.078 |
| **Female (%)** | 4740 (47.8) | 1202 (45.6) | 0.044 | 2156 (45.6) | 996 (45.1) | 0.009 |
| **Race/Ethnicity (%)** | | | 0.700 | | | 0.032 |
| Non-Hispanic White | 5628 (56.8) | 711 (27.0) | | 1951 (41.3) | 698 (31.6) | |
| Non-Hispanic Black | 1091 (11.0) | 222 (8.4) | | 463 (9.8) | 197 (8.9) | |
| Hispanic | 2114 (21.3) | 1146 (43.5) | | 1593 (33.7) | 834 (37.8) | |
| Other | 1073 (10.8) | 555 (21.1) | | 720 (15.2) | 477 (21.6) | |
| ***Child's parent/household characteristics*** | | | | | | |
| **Parent Education (%)** | | | 0.531 | | | 0.054 |
| Less High School | 623 (6.3) | 603 (22.9) | | 411 (8.7) | 285 (12.9) | |
| High School Graduate or Equivalent | 2383 (24.1) | 605 (23.0) | | 1114 (23.6) | 542 (24.6) | |
| Some College or Associate Degree | 3045 (30.7) | 461 (17.5) | | 1141 (24.1) | 450 (20.4) | |
| Bachelor's Degree or Higher | 3855 (38.9) | 965 (36.6) | | 2061 (43.6) | 929 (42.1) | |
| **Parent Employment (%)** | 3211 (32.4) | 715 (27.1) | 0.115 | 1367 (28.9) | 620 (28.1) | 0.018 |
| **Household Poverty (%)** | | | 0.224 | | | 0.091 |
| <1.00 | 1080 (10.9) | 424 (16.1) | | 521 (11.0) | 299 (13.6) | |
| 1.00–1.99 | 1791 (18.1) | 608 (23.1) | | 914 (19.3) | 454 (20.6) | |
| 2.00–3.99 | 2917 (29.4) | 705 (26.8) | | 1366 (28.9) | 612 (27.7) | |
| 4.00+ | 4118 (41.6) | 897 (34.1) | | 1926 (40.7) | 841 (38.1) | |
| **Household Structure (%)** | 7113 (71.8) | 2009 (76.3) | 0.102 | 3586 (75.9) | 1681(76.2) | 0.008 |

**Note:** SMD = standardized mean difference. Values <0.1 indicate good balance.

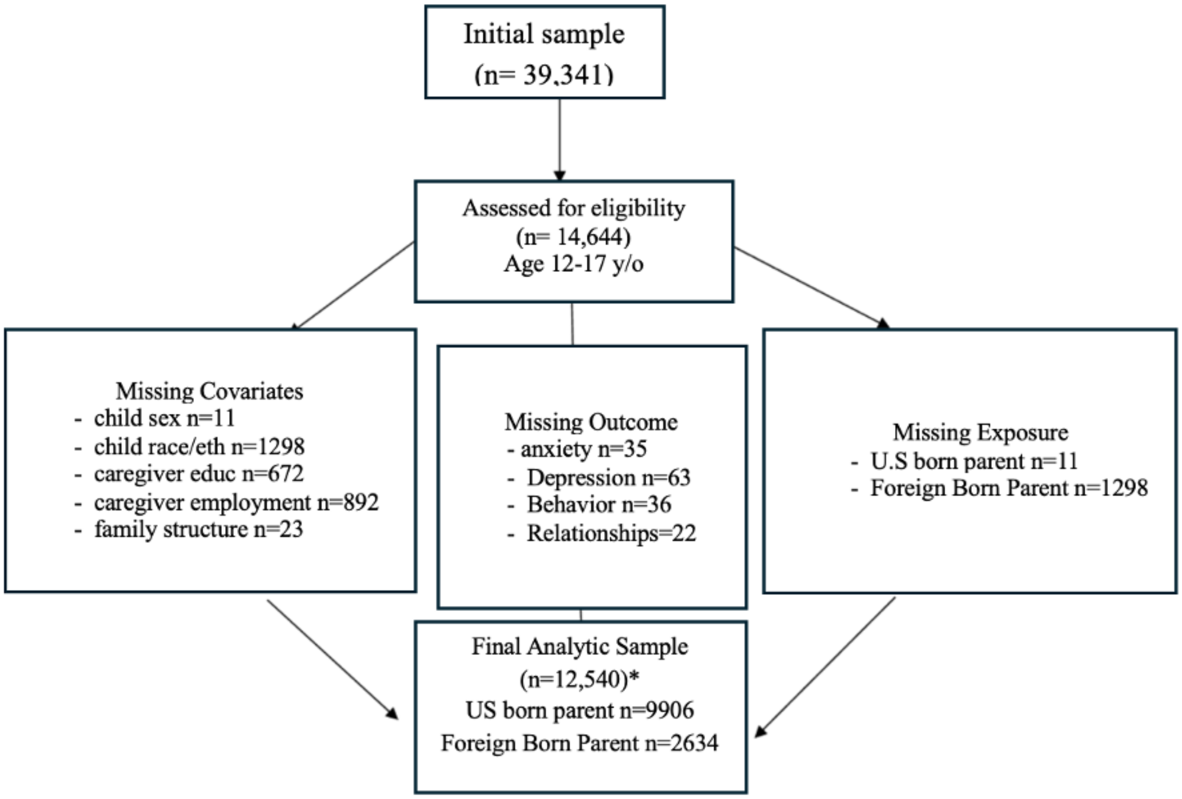


Figure S1: Inclusions and Exclusions for Primary Analytic Sample, NHIS 2022–2024

**Note:** *Exclusion categories are not mutually exclusive; participants may have missing information for more than one variable.

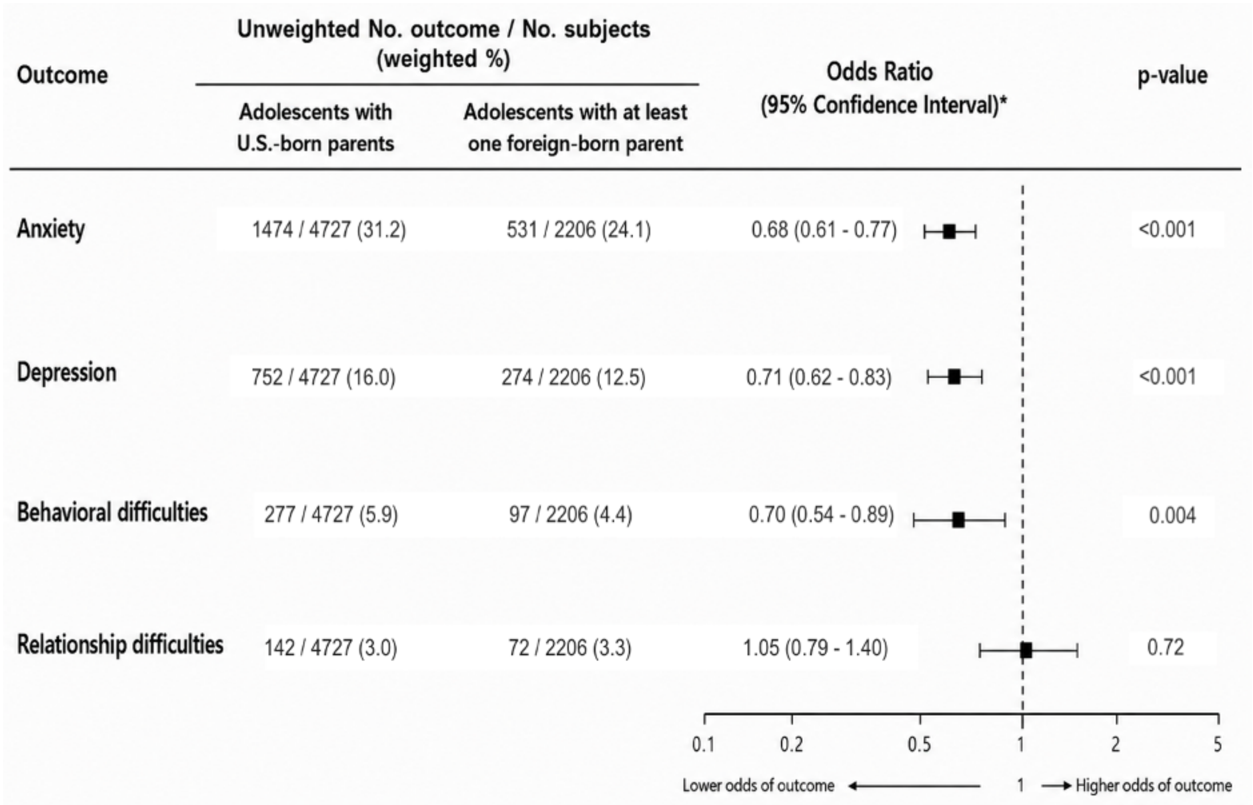

Figure S2: Estimated association between parental nativity and mental health outcomes among adolescents aged 12–17, using nearest-neighbor matching analyses (*denotes the numbers were based on the matched cohort)
